# EM-BENCH: A Reproducible Framework for Benchmarking Guideline-Concordant Clinical Decision Making in Large Language Models

**DOI:** 10.64898/2026.10.27.26366650

**Authors:** Nathan Nguyen, Leland Perice, Rohit B. Sangal, Vishantan Kumar, Andrew Chu, Debadutta Dash

**Affiliations:** Department of Emergency Medicine, Stanford University School of Medicine, Stanford, California; Department of Emergency Medicine, The Warren Alpert Medical School of Brown University, Providence, Rhode Island; Department of Emergency Medicine, Yale School of Medicine, New Haven, Connecticut; Technology & Digital Solutions, Stanford Health Care, Palo Alto, California

**Author notes:** Drs. Chu and Dash contributed equally to this manuscript. **Description:** EM-BENCH is a physician-validated, deterministically scored benchmark of 210 clinical vignettes covering every graded recommendation across the 25 ACEP clinical policies, built as a reproducible template any specialty can use to test large language models for clinical decision support. Across 149 model configurations, every model fell short of full guideline concordance, with performance improving with model size and recency and closed-access models substantially outperforming open-weight models. Corresponding Author: Debadutta Dash, M.D., M.P.H., Department of Emergency Medicine, Stanford University School of Medicine, 900 Welch Road, Suite 350, Palo Alto, CA 94304.

## Abstract

**Background:** Physicians increasingly use artificial intelligence tools to support clinical work, yet there is no accepted standard for measuring the competency of large language models (LLMs) in clinical decision support. We developed EM-BENCH, a physician-validated, deterministically scored benchmark for evaluating whether models reliably apply the American College of Emergency Physicians (ACEP) guidelines to clinical decisions.

**Methods:** EM-BENCH comprises 210 binary clinical vignettes covering all 70 critical questions across 25 ACEP clinical policies. We evaluated 149 LLM configurations representing 79 base models using prespecified answers and deterministic scoring. Each vignette had a prespecified correct answer to a binary yes-or-no question based on an ACEP recommendation. We examined associations between accuracy and model access tier (closed-access vs. open-weight models), model size, release date, clinical policy, recommendation evidence level, and reasoning settings.

**Results:** No configuration achieved complete guideline concordance. The highest performing configuration answered 207 of 210 vignettes correctly (98.6%). Closed-access models outperformed open-weight models (median accuracy, 91% vs. 63%; Hodges–Lehmann location shift, 26 percentage points; 99% CI, 20 to 33; adjusted P<0.001). Among open-weight model families, a 10-fold increase in parameter count was associated with a 14-percentage-point increase in accuracy (99% CI, 7.7 to 20; adjusted P<0.001). Across all models, accuracy increased by 8.9 percentage points per release year (99% CI, 3.6 to 14; adjusted P<0.001). Neither enabling a thinking mode (pseudomedian change, 0.95 percentage points; 99% CI, −2.6 to 3.1; adjusted P=1.00) nor high versus low reasoning effort (0.91 percentage points; 99% CI, −0.69 to 3.1; adjusted P=0.56) consistently improved accuracy.

**Conclusions:** Contemporary LLMs showed improving but incomplete ability to apply ACEP guidelines. Concordance varied markedly across models and was associated with access tier, size, and release recency, while additional inference-time reasoning offered inconsistent benefit. Specialty-specific, physician-validated benchmarks may provide a reproducible assessment of LLMs’ ability to make guideline-concordant clinical decisions.

## Introduction

In a recent *Science* study, Brodeur et al. found that frontier large language models (LLMs) outperform physician comparators on several clinical reasoning tasks.^1^ Their emergency department (ED) evaluation centered on differential diagnosis, although the authors noted that emergency care often depends on “triage, disposition, and immediate management and not diagnostic accuracy.”^1^ A model that correctly diagnoses but fails to suggest appropriate clinical action is not ready for deployment. Rigorous evaluation of LLM clinical decision-making capabilities therefore remains an unmet critical need.

This need is increasingly urgent as physicians incorporate AI into clinical work. In a 2026 American Medical Association survey, 81% of nearly 1,700 physicians reported using artificial intelligence tools professionally.^2^ Reported use for discharge instructions, care plans, or progress notes was nearly twice as common as use for diagnosis. The most frequently reported application was summarizing medical research and standards of care. At the same time, 88% of physicians considered robust safety and efficacy validation critical to adoption.^2^ These findings highlight the need for benchmarks that assess whether models can make appropriate clinical management decisions that align with clinical guidelines.

Specialty guidelines provide a foundation for such evaluation translating systematic evidence review and expert consensus into nationally recognized practice recommendations.³ Existing benchmarks test various skills from medical exam questions to clinical safety assessments.^4–9^ Though they represent important progress, evaluations that use physician behavior or expert consensus as reference standards must contend with variation among clinicians, institutions, and region. This can make the correct clinical decision difficult to adjudicate reproducibly and may encode local practice patterns rather than optimal care. For example, a study grading LLM recommendations against 10,000 real emergency department visits acknowledged that actual patient outcomes may not represent best-practice care.^10^ By grounding evaluation in evidence-based recommendations, specialty guidelines provide a defensible and reproducible baseline for assessing an LLM’s clinical capabilities.

AMEGA and MedGUIDE represent advances in guideline-based evaluation but rely on LLMs in their evaluation pipelines. AMEGA uses physician-developed cases but relies on GPT-4 to score open-ended responses.¹¹ MedGUIDE derives items from National Comprehensive Cancer Network decision trees while using LLMs to generate clinical scenarios and LLM judges to select them.¹² These approaches enable impressive scale and flexibility, but they make the evaluation itself a complex system whose behavior must, in turn, be validated. A 2025 NEJM AI editorial cautioned that benchmarking approaches relying heavily on AI, “if not tethered to extensive human validation, can veer from clinical reality to a machine simulacrum.”¹³ Thus, a benchmark designed for physician trust should have simple machinery: every question physician-validated and no model involved in scoring model responses. There remains a need for benchmarks that meet these criteria while comprehensively representing published recommendations and clinically realistic scenarios.

We therefore developed a reproducible framework for evaluating LLM guideline application, beginning with emergency medicine. The setting is particularly consequential, as emergency departments account for nearly half of U.S. hospital admissions and require time-sensitive decisions for high-acuity presentations.^14^ We created EM-BENCH, a benchmark of 210 physician-reviewed vignettes covering all 70 critical questions – and all 93 associated recommendations – across the 25 American College of Emergency Physicians (ACEP) clinical policies. Each vignette isolates a patient management decision and is scored against a prespecified answer without an evaluator LLM. We evaluated 149 configurations representing 79 base models to examine variation in guideline concordance across various model and policy characteristics.

## Methods

### Benchmark Development and Physician Validation

EM-BENCH consists of 210 clinical vignettes spanning all 25 ACEP clinical policies as of May 31, 2026.^15–39^ The included policies address 70 critical questions, each supported by one or more recommendations graded as Level A, B, or C. Levels A, B, and C reflect high, moderate, and limited clinical certainty, respectively. Each critical question was represented by two to four vignettes (median, three), covering all 93 graded recommendations. The final benchmark included 8 vignettes based on Level A recommendations, 81 based on Level B recommendations, and 121 based on Level C recommendations.

Claude Opus 4.5 and 4.6 were used during initial brainstorming to generate patient presentations relevant to the ACEP recommendations. All 210 initial vignettes were drafted by specifying key details such as clinical history, examination/laboratory findings, and vital signs. Each vignette concluded with a binary yes-or-no question and a prespecified answer derived from the corresponding ACEP recommendation. All vignettes were independently reviewed by three board-certified emergency physicians from two academic medical centers. Reviewers assessed clinical realism, consistency with the cited recommendation, and whether the presented information supported the prespecified answer. Physician review resulted in substantial revision of the initial vignettes, and disagreements were resolved through consensus.

### Model Selection and Configurations

We evaluated 149 model configurations (a base model plus an inference setting) between October 14 and 23, 2026, representing 79 base models from nine developers with release dates spanning December 2023 to October 2026 (Table 1). Closed-access models were accessed through an institutional LiteLLM gateway. Open-weight models were served either locally by Ollama at its default quantization or through Databricks model endpoints, with the same pipeline and prompts on every host (Table S1). Parameter counts for open-weight models were based on the served weights; counts for closed-access models are not published and were excluded from model-size analyses.

**Table 1.**
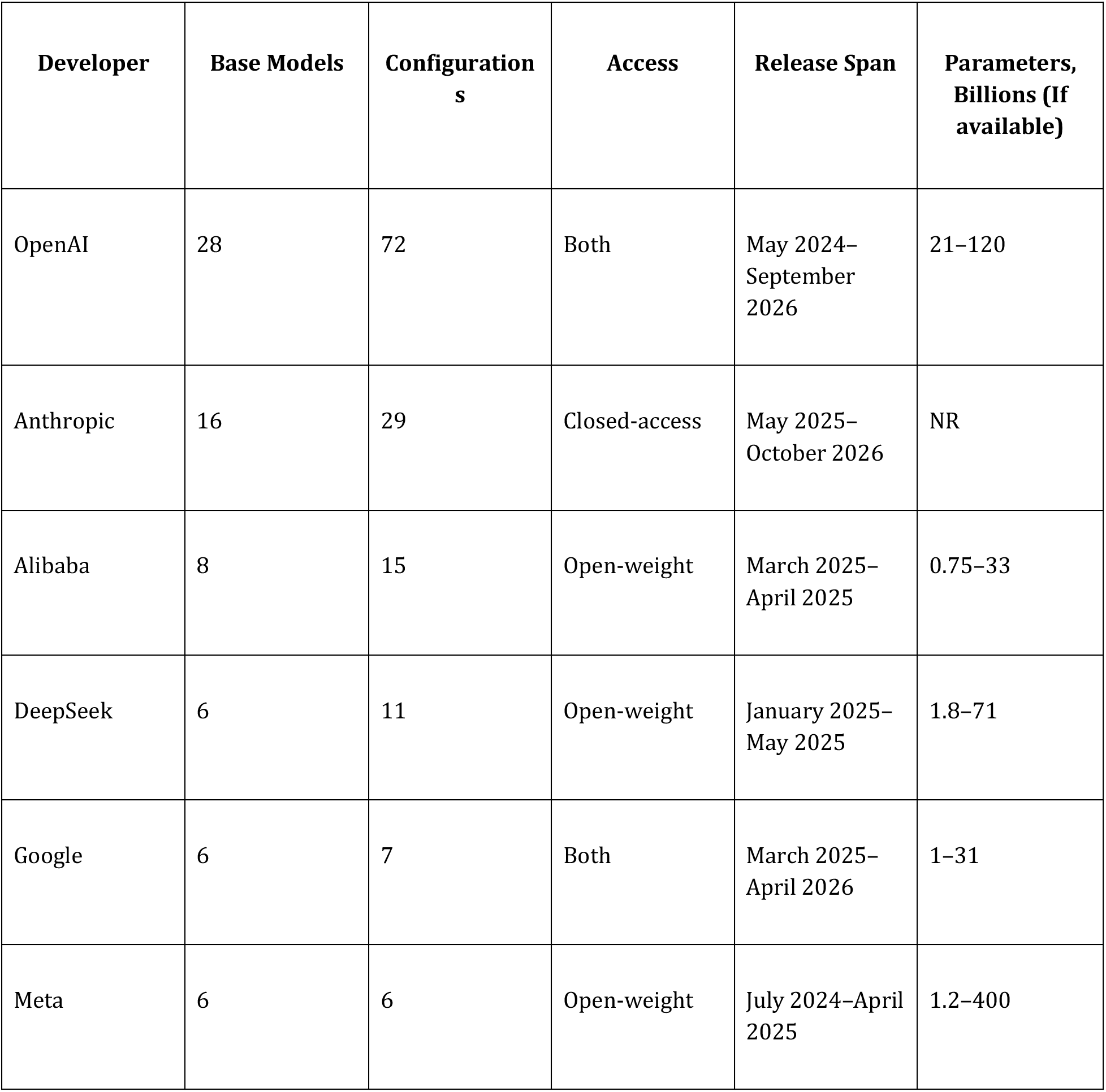
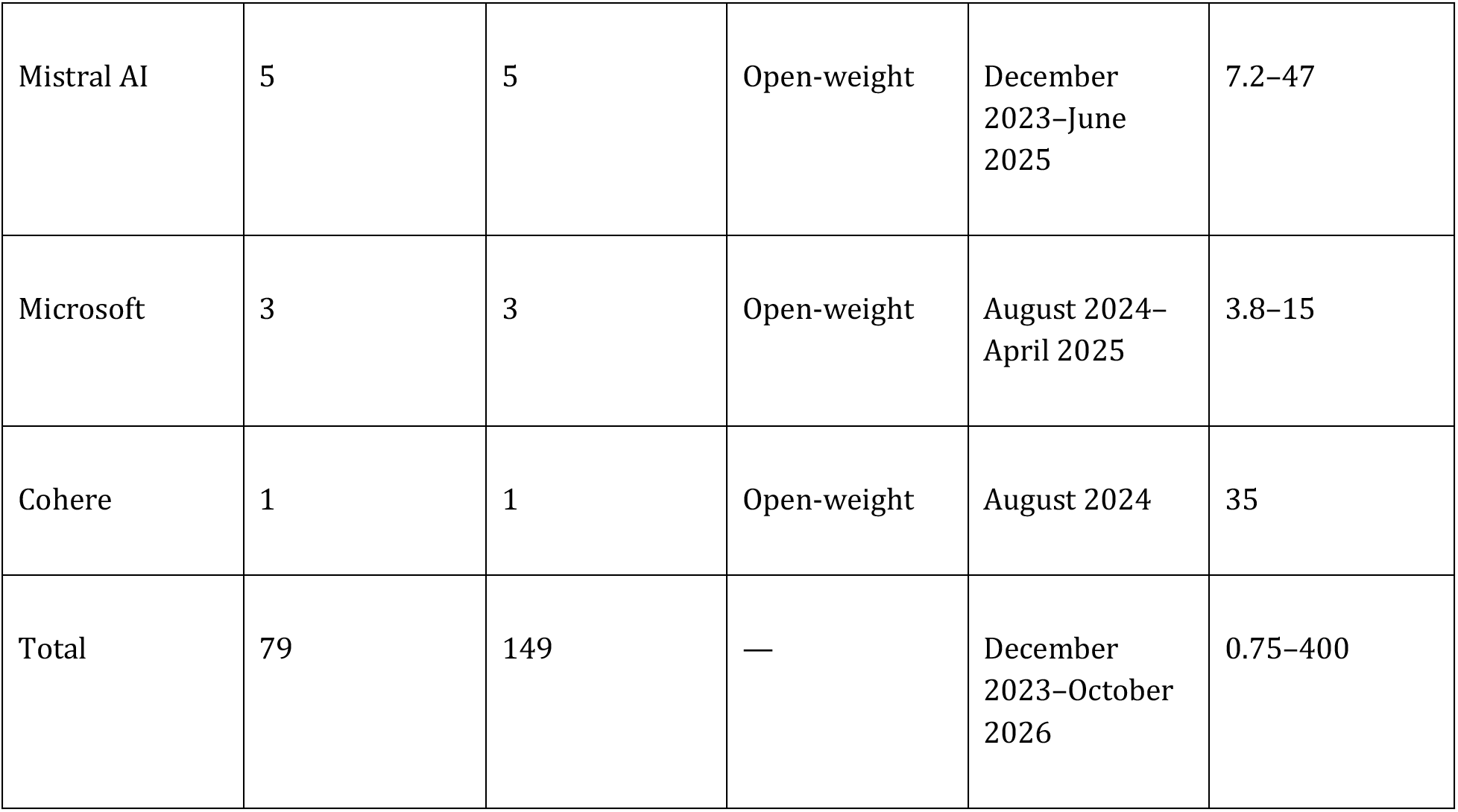
Characteristics of the Large Language Model Panel evaluated in EM-BENCH. Models are summarized by developer, access tier, release span, and, for open-weight models, parameter count of the served weights. Base models with multiple reasoning or thinking settings contributed more than one evaluated configuration. OpenAI’s open-weight models belonged to the gpt-oss family, and Google’s belonged to the Gemma family. NR denotes not reported. Source: EM-BENCH evaluation runs, October 14 to 23, 2026; release dates and parameter counts from developer documentation.

| **Developer** | **Base Models** | **Configurations** | **Access** | **Release Span** | **Parameters, Billions (If available)** |
| --- | --- | --- | --- | --- | --- |
| OpenAI | 28 | 72 | Both | May 2024–September 2026 | 21–120 |
| Anthropic | 16 | 29 | Closed-access | May 2025–October 2026 | NR |
| Alibaba | 8 | 15 | Open-weight | March 2025–April 2025 | 0.75–33 |
| DeepSeek | 6 | 11 | Open-weight | January 2025–May 2025 | 1.8–71 |
| Google | 6 | 7 | Both | March 2025–April 2026 | 1–31 |
| Meta | 6 | 6 | Open-weight | July 2024–April 2025 | 1.2–400 |
| Mistral AI | 5 | 5 | Open-weight | December 2023–June 2025 | 7.2–47 |
| Microsoft | 3 | 3 | Open-weight | August 2024–April 2025 | 3.8–15 |
| Cohere | 1 | 1 | Open-weight | August 2024 | 35 |
| Total | 79 | 149 | — | December 2023–October 2026 | 0.75–400 |

OpenAI reasoning models were evaluated at low, medium, and high reasoning effort, and Claude and compatible open-weight models with thinking enabled and disabled. Claude thinking configurations used each model’s native extended-thinking parameters with the same system prompt as all other configurations. Configurations in which any vignette lacked a valid answer were excluded from the study (Table S1).

### Evaluation Pipeline

Responses were scored deterministically without an evaluator LLM. Models were required to return a single JSON object containing one “answer” key with the value “yes” or “no.” Structured output schemas enforced this format where supported; otherwise, a tiered parser accepted only valid JSON objects. A response was scored correct only if the returned value exactly matched the prespecified answer, so free text and reasoning content were never scored.

Configurations ran at temperature 0 where supported; reasoning-active open-weight configurations used their model cards’ recommended sampling parameters, and endpoints that reject a temperature parameter ran at their defaults. Generation budgets were set to be non-binding, and all scored configurations completed all 210 vignettes without exhausting them. Sampling parameters, schema use, and token budgets for every configuration are provided in Table S1.

### Statistical Analysis

Analyses examined performance by access tier, model size, release date, clinical policy, recommendation level, and reasoning setting. Except for paired reasoning comparisons, accuracy was averaged across configurations so that each base model contributed once. As sample size was determined by guideline coverage and model availability, no prospective power calculation was performed. Performance was summarized by the distribution of base-model accuracy. Closed-access and open-weight models were compared using a two-sided Wilcoxon rank-sum test, with the Hodges–Lehmann location shift and Cliff’s delta as effect measures.

Among open-weight families with at least three sizes, the association between accuracy and log10 total parameter count was estimated using a linear mixed-effects model with a random intercept for model family. Release date was modeled in years using linear regression adjusted for access tier, with standard errors clustered by model lineage. An access-by-release-date interaction was examined as a secondary analysis. Policy specific accuracies were summarized descriptively. Recommendation level accuracy was summarized separately within each access tier. Within closed-access and open-weight models, paired A-minus-C and B-minus-C accuracy differences were estimated using two-sided Wilcoxon signed-rank procedures and reported with unadjusted 95% confidence intervals without P values.

Reasoning effects were evaluated within base models using two-sided Wilcoxon signed-rank tests, only including base models evaluated under both relevant settings. Model-specific exact McNemar tests were exploratory and were Holm-adjusted. The five primary comparisons—access tier, model size, release date, thinking enabled versus disabled, and high versus low reasoning effort—formed one family. Bonferroni-adjusted two-sided P values were evaluated at a familywise significance level of 0.05, with corresponding 99% confidence intervals. Secondary analyses were reported with unadjusted 95% confidence intervals and generally without P values. All included configurations completed all 210 vignettes without item-level missingness, so imputation was not applicable. Analyses were performed using R, version 4.5.2 (R Foundation for Statistical Computing), through RStudio, version 2026.01.0+392 (Posit Software), with the tidyverse, lme4, lmerTest, and sandwich packages.

## Results

### Overall Concordance and Differences by Access Tier

No evaluated configuration achieved complete guideline concordance, and performance separated sharply by access tier. GPT-6 Astra at high reasoning effort was the best performing configuration, answering 207 of 210 vignettes correctly (98.6%). Qwen3 0.6B with thinking enabled was the worst performing configuration, answering 79 of 210 correctly (37.6%). Base model accuracy ranged from 41% to 98%, with a median of 83% (Figure 1). Closed-access models (median, 91%) outperformed open-weight models (median, 63%), with a Hodges–Lehmann location shift of 26 percentage points (99% CI, 20 to 33; adjusted P<0.001). Cliff’s delta was 0.96 (99% CI, 0.87 to 1.00), indicating near-complete separation between access tiers.

**Figure 1.**
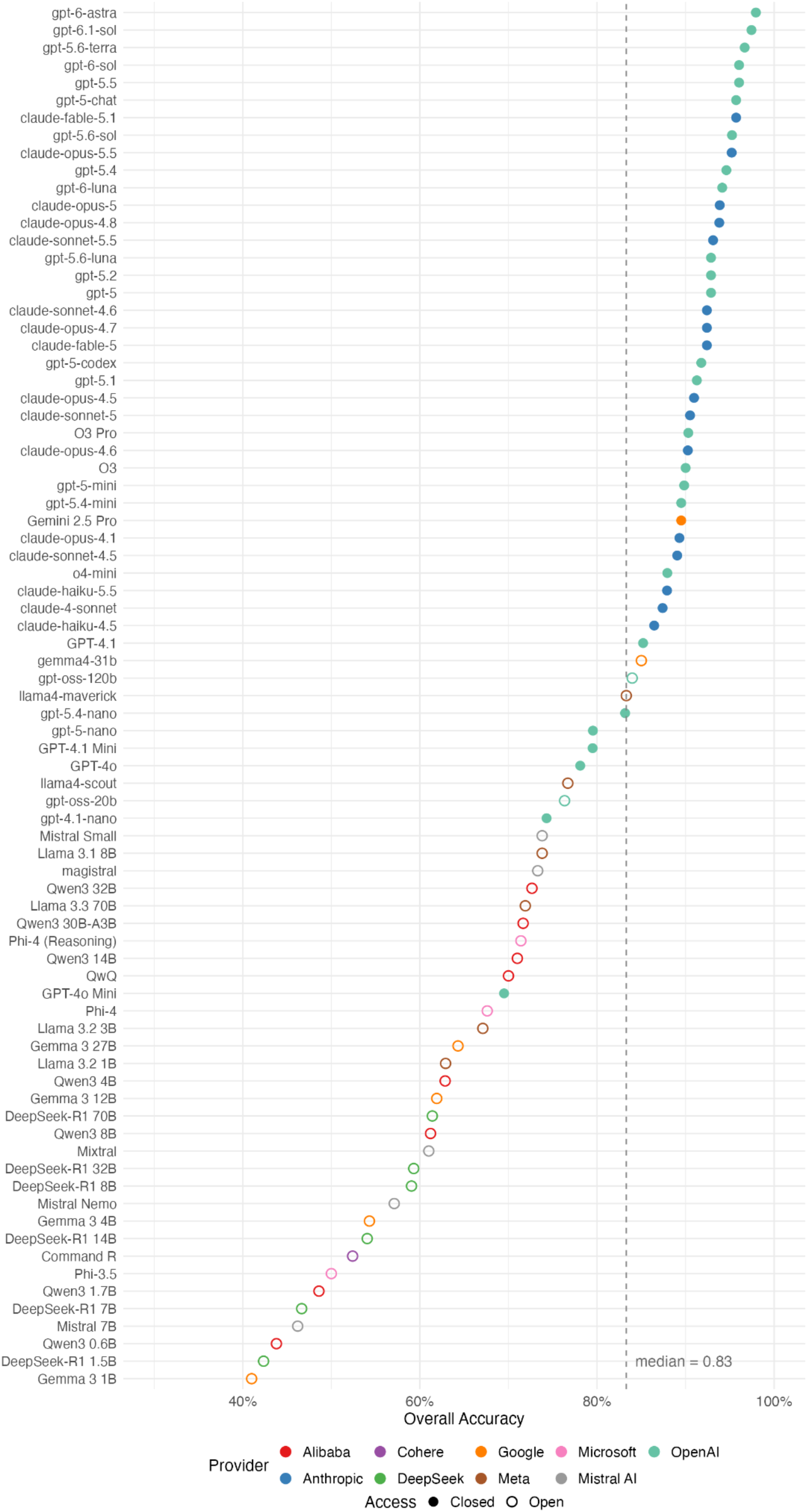
Accuracy Across Evaluated Large Language Models. Overall accuracy for each base model on the 210-vignette EM-BENCH benchmark, ranked from highest to lowest. Accuracy was averaged across inference configurations when multiple configurations of the same base model were evaluated. Colors indicate model developer; filled and hollow circles indicate closed-access and open-weight models, respectively. The dashed vertical line indicates the median accuracy across base models. Source: authors’ analysis of EM-BENCH results.

### Accuracy by Model Size and Release Recency

Accuracy increased consistently with model size and release recency. Among 28 base models in six open-weight families, a 10-fold increase in parameter count was associated with a 14-percentage-point increase in accuracy (99% CI, 7.7 to 20; adjusted P<0.001) (Figure 2A). The association was positive in all six families, with within-family Spearman correlations ranging from 0.80 to 1.00. Across all 79 base models, accuracy increased by 8.9 percentage points per release year (99% CI, 3.6 to 14; adjusted P<0.001) (Figure 2B). The estimated difference between the tier-specific temporal trends was 4.8 percentage points per year (95% CI, −5.6 to 15).

**Figure 2.**
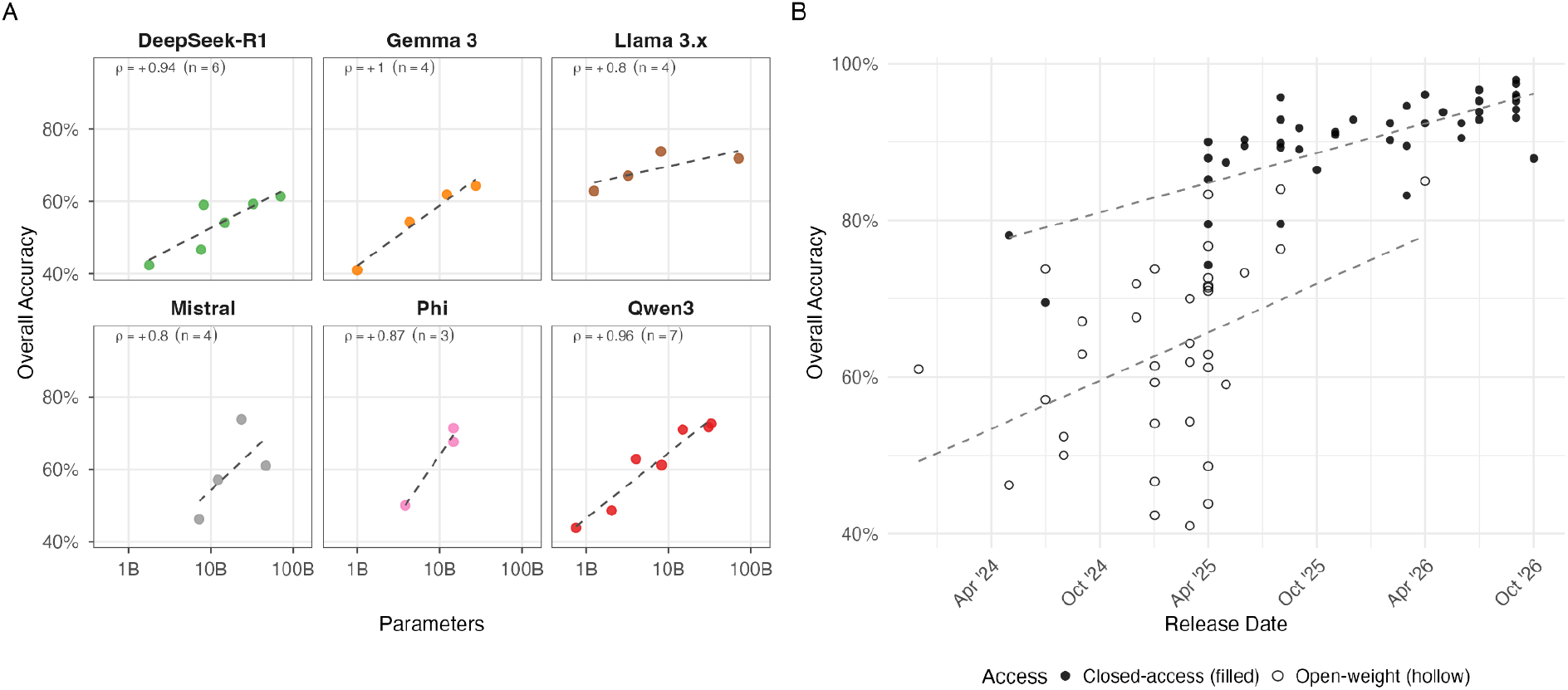
Associations of Model Size and Release Date With Guideline Concordance. Panel A shows overall accuracy according to parameter count among open-weight model families with at least three evaluated model sizes. Each point represents a base model; dashed lines show within-family linear fits, and Spearman correlation coefficients (ρ) are shown for each family. Panel B shows overall accuracy according to model release date across closed-access and open-weight models. Filled and hollow circles indicate closed-access and open-weight models, respectively; dashed lines show fitted trends within each access tier. Source: authors’ analysis of EM-BENCH results.

### Accuracy Varied by Policy and Was Lowest on Level A Recommendations

Performance varied widely across clinical policies, and both access tiers were least accurate on Level A recommendations. Pooled policy accuracy ranged from 58% (95% CI, 54 to 63) for seizures to 92% (95% CI, 91 to 94) for adult psychiatric care (Figure 3A). Mean accuracy among closed-access models was 81%, 91%, and 90% for Level A, B, and C recommendations, respectively; the paired differences were −9.7 percentage points for A versus C (95% CI, −11 to −6.8) and 1.2 percentage points for B versus C (95% CI, 0.47 to 2.5). Mean open-weight accuracy was 47%, 60%, and 66%; the paired differences were −18 percentage points for A versus C (95% CI, −24 to −13) and −5.2 percentage points for B versus C (95% CI, −7.9 to −3.9) (Figure 3B). No base model answered all eight Level A vignettes correctly.

**Figure 3.**
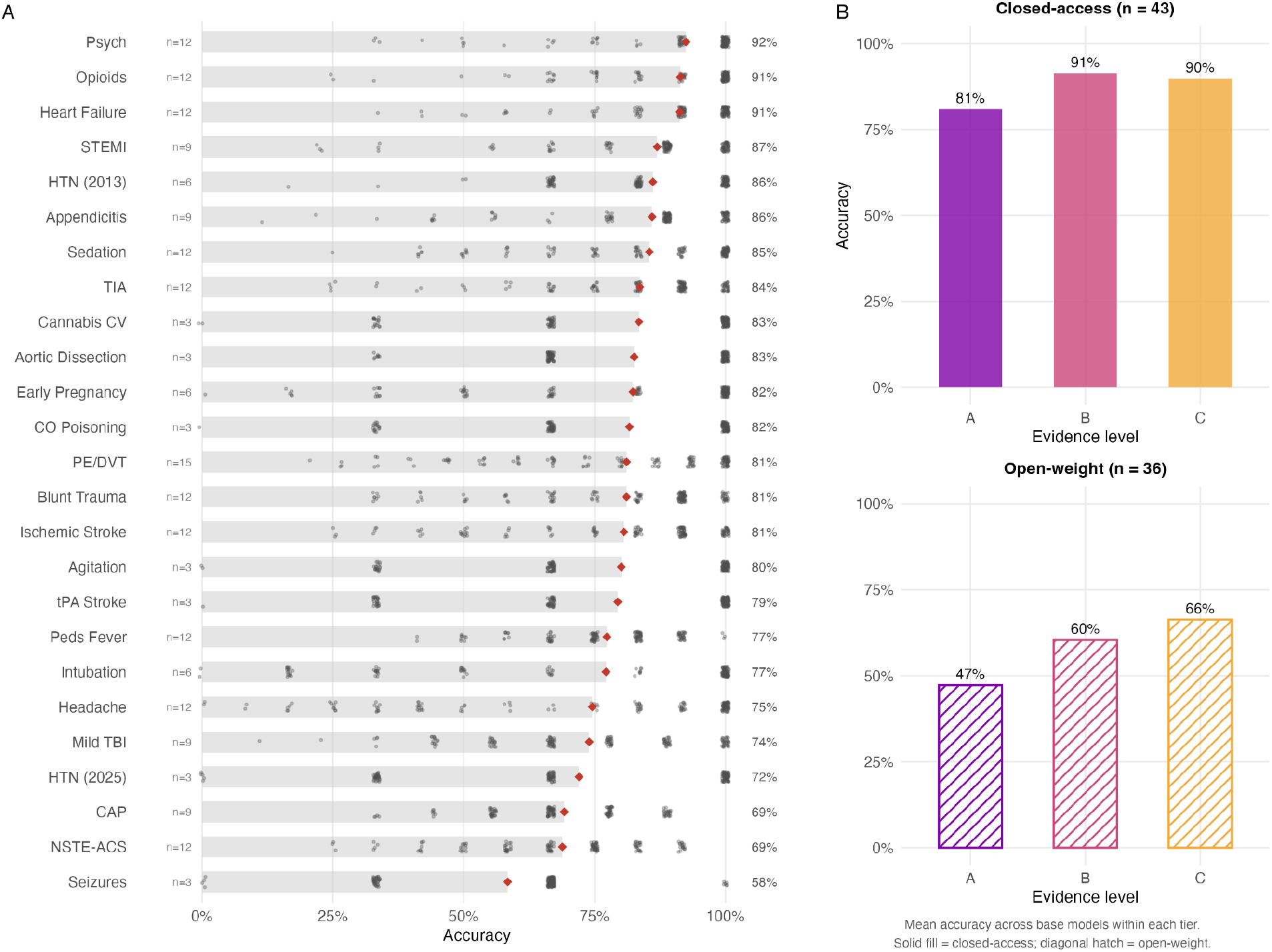
Guideline Concordance Across Clinical Policies and Recommendation Evidence Levels. Panel A shows accuracy across the 25 evaluated ACEP clinical policies. Gray points represent individual model-configuration accuracies within each policy, red diamonds indicate pooled accuracy, and n denotes the number of unique vignettes representing that policy. Panel B shows mean accuracy across base models for Level A, B, and C recommendations, stratified by access tier. EM-BENCH included 8 Level A, 81 Level B, and 121 Level C vignettes. Source: authors’ analysis of EM-BENCH results.

### Reasoning Settings Did Not Improve Accuracy Overall but Varied Across Model Families

Neither reasoning setting improved accuracy overall. Across 26 paired models, the mean change with thinking enabled was 0.22 percentage points, and the Hodges–Lehmann pseudomedian change was 0.95 percentage points (99% CI, −2.6 to 3.1; adjusted P=1.00) (Figure 4). Accuracy increased in 17 models, decreased in 8, and was unchanged in 1. Changes ranged from a 12.4-percentage-point decrease for Qwen3 0.6B to a 7.1-percentage-point increase for DeepSeek-R1 14B. Among 22 OpenAI models evaluated at both high and low reasoning effort, the pseudomedian change was 0.91 percentage points (99% CI, −0.69 to 3.1; adjusted P=0.56), with improvement in 12 of 22 models. Additional reasoning improved accuracy in 11 of 13 Claude models with thinking enabled and in both gpt-oss models at medium and high effort, with inconsistent effects across other model families; no model-specific exact McNemar test was significant after Holm adjustment.

**Figure 4.**
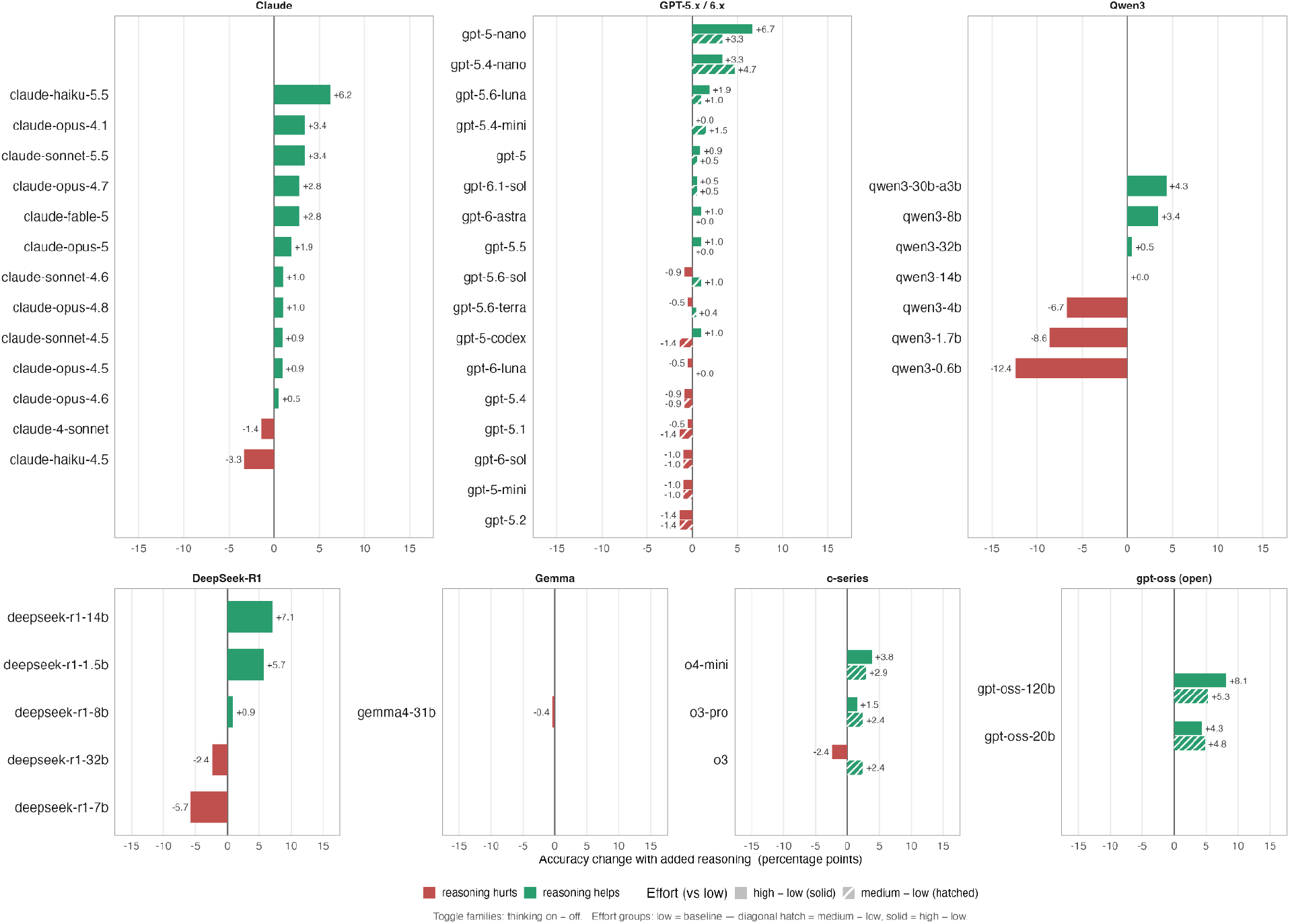
Effect of Additional Reasoning on Guideline Concordance. Accuracy changes associated with additional inference-time reasoning are shown for individual base models. For models evaluated with a thinking toggle, bars represent the percentage-point difference between thinking-enabled and thinking-disabled configurations. For models evaluated at multiple reasoning-effort levels, hatched and solid bars represent the differences between medium and low effort and between high and low effort, respectively. Positive values indicate higher accuracy with additional reasoning; negative values indicate lower accuracy. Source: authors’ analysis of EM-BENCH results.

## Discussion

None of the LLM configurations achieved complete concordance with the 25 ACEP clinical policies as assessed in EM-BENCH. Accuracy ranged from 37.6% to 98.6%. Closed-access models had higher concordance than open-weight models, and accuracy was positively correlated with parameter count within model families and with release recency. Inference-time reasoning did not improve concordance overall, although effects varied by implementation: most Claude models gained modestly with thinking enabled, and the open-weight gpt-oss models gained at every effort level.

EM-BENCH enters a field actively determining how AI should be evaluated to earn its place in clinical environments. As LLMs have saturated examination-style assessments, evaluation has shifted toward increasingly complex alternatives, many being highly engineered systems that use LLMs themselves across multiple stages of the benchmark workflow to generate questions or judge open-ended responses. Yet Rodman et al. warn that such approaches carry “dubious validity” when untethered from “extensive human validation,” and support a progression from human-grounded assessments to eventual clinical trials.^13^ EM-BENCH creates a foundational checkpoint for such a human-grounded approach. Its vignettes are based on trusted guidelines formed from extensive literature review and expert discussion,^40^ validated by board-certified emergency physicians, and deterministically scored without judge LLMs whose own behavior requires validation.

Related human-grounded benchmarks measure complementary constructs. McCoy et al. tested how models revise clinical judgments as new information arrives, using 750 physician-validated questions adapted from script-concordance tests.^41^ MedAgentBench evaluated retrieval and action on physician-written tasks in a virtual electronic health record.^42^ Within this landscape, EM-BENCH assesses guideline knowledge and application across all 25 ACEP clinical policies. EM-BENCH’s predefined scoring also carries practical weight: any new model can be evaluated without repeated physician adjudication of their responses, and vignettes can and should be updated as policies evolve. Evaluations requiring physician adjudication, while valuable, can be time and labor intensive, making it difficult to keep up with rapid model releases. This design enabled this study to evaluate a model panel several times the size of similar studies, broad enough to support the statistical associations examined.^11,42^

Our findings contribute to an ongoing discussion over whether open-weight models match closed-access model performance across clinical tasks. On complex diagnostic cases drawn from published clinicopathological conferences, the open-weight Llama 3.1 405B performed comparably to GPT-4, and AMEGA’s strongest open-weight and closed-access models scored within a point of each other.^11,43^ However, MedAgentBench reported a gap between the access tiers, which EM-BENCH supports with regard to guideline concordance, as the median open-weight model answered fewer than two of every three vignettes correctly.^42^ This substantial shortfall raises concerns about relying on these models for guideline-based decision support, even where privacy and institutional control favor their deployment.^44^ Notably, the best open-weight models in this study were the largest or most recent available, and accuracy rose with parameter count across the six open-weight families. Accuracy also rose with release recency in both access tiers, although these findings do not establish that the gap between them is narrowing.

The mixed effects of inference-time reasoning are consistent with prior work. McCoy et al. suggested that chain-of-thought reasoning may hinder clinical judgment in uncertain conditions, while Oh et al. found that additional inference-time computation benefits medical tasks unevenly across models and task types.^41,45^ One plausible explanation is that longer reasoning does not fully compensate for gaps in underlying guideline knowledge, consistent with work identifying medical knowledge as a constraint on further reasoning gains.^46^ The exploratory pattern of lower accuracy on Level A recommendations in both access tiers rests on eight vignettes, owing to the scarcity of Level A recommendations across the ACEP clinical policies, and warrants further investigation.

Encouragingly, the most advanced models are approaching complete concordance on EM-BENCH, demonstrating that such LLMs can recall and apply ACEP guidance with impressive, though still incomplete, consistency. We propose complete concordance as a reasonable foundational expectation for two reasons. First, ACEP clinical policies address the most urgent questions in emergency medicine and are among the most discussed and applied guidelines in the specialty.^40^ Given that physicians already use AI to reference standards of care and create care plans, models supporting these tasks should, at a minimum, reliably apply such recommendations.^2^ Second, every answer is prespecified from a published recommendation, and every vignette supplies the information required to adhere to ACEP guidelines. Under these favorable conditions, a miss represents a departure from systematically developed, evidence-based or consensus-backed ACEP recommendations for the clinical scenario presented. The leading configuration missed three vignettes, each answered correctly by several other configurations and attributable to a specific area of emergency medicine care. Given the consequences of decisions in emergency care, these gaps warrant targeted correction as part of preparation for patient-facing use. We propose concordance with ACEP’s recommendations in clearly specified scenarios as a foundational floor before models tackle more complex, uncertain, and high-acuity clinical decisions. Furthermore, frontier LLMs are not necessarily the models that will be deployed in practice. Cost and logistical constraints may favor deployment of other models, whose shortcomings can be obscured when frontier performance defines the field.^44^

Our broader aim is to support a cross-specialty effort to establish a shared baseline for clinical AI evaluation. EM-BENCH offers a reproducible framework through which specialties can translate their evidence-graded guidelines into physician-validated benchmarks. Studies of human–computer interaction and prospective clinical outcomes remain essential to establish clinical readiness.^13^ As in clinical training, confidence can build through progressive assessment of defined competencies. A shared foundation of demonstrated guideline knowledge and application would give this process a transparent starting point, grounding clinician trust in capabilities that have been tested directly.

## Limitations

This study has several limitations. First, EM-BENCH was designed as a constrained test of guideline application rather than a simulation of clinical practice. Its simplified structure enables deterministic evaluation, but real encounters can be significantly more complex. For these reasons, performance on EM-BENCH cannot be extrapolated to real-world performance. Second, only eight vignettes represented Level A recommendations, limiting interpretation of differences across evidence tiers. Third, aggregate accuracy weighted all model incorrect answers equally, although their potential clinical consequences may differ. Fourth, no physician comparator was included by design because the study assessed LLM concordance with published guidance rather than performance relative to clinicians. These findings therefore do not establish whether models outperform physicians or improve clinician decisions. Fifth, translating certain recommendations, particularly Level C guidance, into binary answers requires interpretation and may not capture the full flexibility of the source policies.

## Conclusion

None of the 149 evaluated configurations achieved complete concordance with ACEP recommendations, although the strongest approached it. Performance varied by access tier, model size, and release recency, while additional inference-time reasoning offered inconsistent benefit. EM-BENCH provides a transparent, physician-validated assessment of guideline knowledge and application, a foundational capability that should be demonstrated rather than assumed. Extending this framework across specialties could establish a shared baseline for clinical AI evaluation and contribute to a staged, human-grounded body of evidence for physician trust. Demonstrating this capability would represent progress toward clinical readiness, which still requires evaluation within the intended clinical use.

## Ethics and IRB determination

This study evaluated LLM outputs using synthetic clinical vignettes and did not involve human participants, patient-level data, or identifiable private information. Institutional review board review and informed consent were therefore not applicable.

## AI Use Statement

Use of Artificial Intelligence–Assisted Technologies: Claude Opus 4.5 and Claude Opus 4.6 (Anthropic) were used to brainstorm candidate clinical vignette concepts during benchmark development. GPT-5.6 Sol (OpenAI), Claude Opus 4.6, and Claude Fable 5.1 (Anthropic) were used for editorial and formatting assistance during manuscript preparation. GPT-5.6 Sol, Claude Opus 4.6, Claude Fable 5, and Claude Fable 5.1 assisted with construction of the evaluation and analysis pipelines, as well as with sourcing model metadata. All AI-assisted material was reviewed, verified against the relevant source materials, and substantively edited by the authors. The authors take responsibility for the accuracy, originality, and integrity of the submitted work; AI-assisted technologies were not used as authors or cited as primary sources.

## Data Sharing Statement

The benchmark vignettes with prespecified answers, the exact prompts and output schemas, and per-configuration results will be made available as supplementary material upon publication of the peer-reviewed article. The evaluation and analysis code will be made publicly available in a repository upon publication.

## References

1. Brodeur PG, Buckley TA, Kanjee Z, et al. Performance of a large language model on the reasoning tasks of a physician. Science 2026;392:524–527. DOI: 10.1126/science.adz4433.

2. American Medical Association. More than 80% of physicians use AI professionally: AMA survey. March 12, 2026 (https://www.ama-assn.org/practice-management/digital-health/more-80-physicians-use-ai-professionally-ama-survey).

3. Institute of Medicine. Clinical practice guidelines we can trust. Washington, DC: National Academies Press, 2011. DOI: 10.17226/13058.

4. Jin D, Pan E, Oufattole N, Weng WH, Fang H, Szolovits P. What disease does this patient have? A large-scale open domain question answering dataset from medical exams. Appl Sci 2021;11:6421. DOI: 10.3390/app11146421.

5. Arora RK, Wei J, Soskin Hicks R, et al. HealthBench: evaluating large language models towards improved human health. arXiv. Preprint posted May 13, 2025. DOI: 10.48550/arXiv.2505.08775.

6. Tan A, Dai S, Wang J, et al. A decade-scale benchmark evaluating LLMs’ clinical practice guidelines detection and adherence in multi-turn conversations. arXiv. Preprint posted March 26, 2026. DOI: 10.48550/arXiv.2603.25196.

7. Wang S, Tang Z, Yang H, et al. A novel evaluation benchmark for medical LLMs illuminating safety and effectiveness in clinical domains. NPJ Digit Med 2026;9:91. DOI: 10.1038/s41746-025-02277-8.

8. Mehandru N, Golchini N, Garg N, et al. ER-Reason: a benchmark dataset for LLM clinical reasoning in the emergency room. arXiv. Preprint posted May 28, 2025; revised May 11, 2026. DOI: 10.48550/arXiv.2505.22919.

9. Nedos I, Zagalioti SC, Kofos C, et al. Is artificial intelligence ready for emergency department triage? A retrospective evaluation of multiple large language models in 39,375 patients at a university emergency department. J Clin Med 2026;15:1512. DOI: 10.3390/jcm15041512.

10. Williams CYK, Miao BY, Kornblith AE, Butte AJ. Evaluating the use of large language models to provide clinical recommendations in the emergency department. Nat Commun 2024;15:8236. DOI: 10.1038/s41467-024-52415-1.

11. Fast D, Adams LC, Busch F, et al. Autonomous medical evaluation for guideline adherence of large language models. NPJ Digit Med 2024;7:358. DOI: 10.1038/s41746-024-01356-6.

12. Li X, Gao M, Hao Y, et al. MedGUIDE: benchmarking clinical decision-making in large language models. arXiv. Preprint posted May 16, 2025. DOI: 10.48550/arXiv.2505.11613.

13. Rodman A, Zwaan L, Olson A, Manrai AK. When it comes to benchmarks, humans are the only way. NEJM AI 2025;2(4). DOI: 10.1056/AIe2500143.

14. Pines JM, Mutter RL, Zocchi MS. Variation in emergency department admission rates across the United States. Med Care Res Rev 2013;70:218–231. DOI: 10.1177/1077558712470565.

15. Tomaszewski CA, Nestler D, Shah KH, Sudhir A, Brown MD. Clinical policy: critical issues in the evaluation and management of emergency department patients with suspected non–ST-elevation acute coronary syndromes. Ann Emerg Med 2018;72:e65–e106. DOI: 10.1016/j.annemergmed.2018.07.045.

16. Diercks DB, Adkins EJ, Harrison N, Sokolove PE, Kwok H, Wolf SJ. Clinical policy: critical issues in the evaluation and management of emergency department patients with suspected appendicitis. Ann Emerg Med 2023;81:e115–e152. DOI: 10.1016/j.annemergmed.2023.01.015.

17. Gemme S, Meltzer AC, Byyny R, Diercks DB. Clinical policy: a critical issue in the outpatient management of adult patients presenting to the emergency department with asymptomatic elevated blood pressure. Ann Emerg Med 2025;86:e1–e11. DOI: 10.1016/j.annemergmed.2024.09.016.

18. Wolf SJ, Lo B, Shih RD, Smith MD, Fesmire FM. Clinical policy: critical issues in the evaluation and management of adult patients in the emergency department with asymptomatic elevated blood pressure. Ann Emerg Med 2013;62:59–68. DOI: 10.1016/j.annemergmed.2013.05.012.

19. Gerardo CJ, Blanda M, Garg N, et al. Clinical policy: critical issues in the evaluation of adult patients presenting to the emergency department with acute blunt trauma. Ann Emerg Med 2024;84:e25–e55. DOI: 10.1016/j.annemergmed.2024.05.027.

20. Godwin SA, Burton JH, Gerardo CJ, et al. Clinical policy: procedural sedation and analgesia in the emergency department. Ann Emerg Med 2014;63:247–258.e18. DOI: 10.1016/j.annemergmed.2013.10.015.

21. Godwin SA, Hahn SA, Friedman BW, et al. Clinical policy: critical issues in the management of adult patients requiring endotracheal intubation in the emergency department. Ann Emerg Med 2025;86:e29–e68. DOI: 10.1016/j.annemergmed.2025.04.003.

22. Hahn SA, Promes SB, Brown MD. Clinical policy: critical issues in the initial evaluation and management of patients presenting to the emergency department in early pregnancy. Ann Emerg Med 2017;69:241–250.e20. DOI: 10.1016/j.annemergmed.2016.11.002.

23. Hatten BW, Bonney C, Wall SP, Diercks DB. Clinical policy: critical issues related to harms of cannabis exposure in adult patients presenting to the emergency department, cardiovascular considerations. Ann Emerg Med 2026;87:e1–e9. DOI: 10.1016/j.annemergmed.2025.06.608.

24. Lo BM, Carpenter CR, Hatten BW, Wright BJ, Brown MD. Clinical policy: critical issues in the evaluation of adult patients with suspected transient ischemic attack in the emergency department. Ann Emerg Med 2016;68:354–370.e29. DOI: 10.1016/j.annemergmed.2016.06.048.

25. Lo BM, Carpenter CR, Ducey S, Gottlieb M, Kaji A, Diercks DB. Clinical policy: critical issues in the management of adult patients presenting to the emergency department with acute ischemic stroke. Ann Emerg Med 2023;82:e17–e64. DOI: 10.1016/j.annemergmed.2023.03.007.

26. Lo BM, Carpenter CR, Milne K, Panagos P, Haukoos JS, Diercks DB. Clinical policy: use of thrombolytics for the management of acute ischemic stroke in the emergency department. Ann Emerg Med 2024;84:e57–e86. DOI: 10.1016/j.annemergmed.2024.07.023.

27. Mace SE, Gemme SR, Valente JH, et al. Clinical policy for well-appearing infants and children younger than 2 years of age presenting to the emergency department with fever. Ann Emerg Med 2016;67:625–639.e13. DOI: 10.1016/j.annemergmed.2016.01.042.

28. Nazarian DJ, Broder JS, Thiessen MEW, Wilson MP, Zun LS, Brown MD. Clinical policy: critical issues in the diagnosis and management of the adult psychiatric patient in the emergency department. Ann Emerg Med 2017;69:480–498. DOI: 10.1016/j.annemergmed.2017.01.036.

29. Promes SB, Glauser JM, Smith MD, Torbati SS, Brown MD. Clinical policy: emergency department management of patients needing reperfusion therapy for acute ST-segment elevation myocardial infarction. Ann Emerg Med 2017;70:724–739. DOI: 10.1016/j.annemergmed.2017.09.035.

30. Promes SB, Westafer L, Byyny R, Diercks DB. Critical issues in the evaluation and management of adult patients with suspected acute nontraumatic thoracic aortic dissection. Ann Emerg Med 2025;86:e12–e26. DOI: 10.1016/j.annemergmed.2025.04.002.

31. Shih RD, Tomaszewski CA, Kaji A, Diercks DB. A critical issue in the management of adult patients presenting to the emergency department with acute carbon monoxide poisoning. Ann Emerg Med 2025;85:e45–e59. DOI: 10.1016/j.annemergmed.2024.12.005.

32. Silvers SM, Gemme SR, Hickey S, et al. Clinical policy: critical issues in the evaluation and management of adult patients presenting to the emergency department with acute heart failure syndromes. Ann Emerg Med 2022;80:e31–e59. DOI: 10.1016/j.annemergmed.2022.05.027.

33. Smith MD, Sampson CS, Wall SP, Diercks DB. Clinical policy: critical issues in the management of adult patients presenting to the emergency department with seizures. Ann Emerg Med 2024;84:e1–e12. DOI: 10.1016/j.annemergmed.2024.02.018.

34. Thiessen MEW, Godwin SA, Hatten BW, Whittle JA, Haukoos JS, Diercks DB. Clinical policy: critical issues in the evaluation and management of adult out-of-hospital or emergency department patients presenting with severe agitation. Ann Emerg Med 2024;83:e1–e30. DOI: 10.1016/j.annemergmed.2023.09.010.

35. Valente JH, Anderson JD, Paolo WF, et al. Clinical policy: critical issues in the management of adult patients presenting to the emergency department with mild traumatic brain injury. Ann Emerg Med 2023;81:e63–e105. DOI: 10.1016/j.annemergmed.2023.01.014.

36. Wolf SJ, Hahn SA, Nentwich LM, Raja AS, Silvers SM, Brown MD. Clinical policy: critical issues in the evaluation and management of adult patients presenting to the emergency department with suspected acute venous thromboembolic disease. Ann Emerg Med 2018;71:e59–e109. DOI: 10.1016/j.annemergmed.2018.03.006.

37. Godwin SA, Cherkas DS, Panagos PD, Shih RD, Byyny R, Wolf SJ. Clinical policy: critical issues in the evaluation and management of adult patients presenting to the emergency department with acute headache. Ann Emerg Med 2019;74:e41–e74. DOI: 10.1016/j.annemergmed.2019.07.009.

38. Hatten BW, Cantrill SV, Dubin JS, et al. Clinical policy: critical issues related to opioids in adult patients presenting to the emergency department. Ann Emerg Med 2020;76:e13–e39. DOI: 10.1016/j.annemergmed.2020.06.049.

39. Smith MD, Fee C, Mace SE, et al. Clinical policy: critical issues in the management of adult patients presenting to the emergency department with community-acquired pneumonia. Ann Emerg Med 2021;77:e1–e57. DOI: 10.1016/j.annemergmed.2020.10.024.

40. American College of Emergency Physicians. Clinical policies (https://www.acep.org/patient-care/clinical-policies). Accessed October 27, 2026.

41. McCoy LG, Swamy R, Sagar N, et al. Assessment of large language models in clinical reasoning: a novel benchmarking study. NEJM AI 2025;2(10). DOI: 10.1056/AIdbp2500120.

42. Jiang Y, Black KC, Geng G, et al. MedAgentBench: a virtual EHR environment to benchmark medical LLM agents. NEJM AI 2025;2(9). DOI: 10.1056/AIdbp2500144.

43. Buckley TA, Crowe B, Abdulnour RE, Rodman A, Manrai AK. Comparison of frontier open-source and proprietary large language models for complex diagnoses. JAMA Health Forum 2025;6:e250040. DOI: 10.1001/jamahealthforum.2025.0040.

44. Dennstädt F, Hastings J, Putora PM, Schmerder M, Cihoric N. Implementing large language models in healthcare while balancing control, collaboration, costs and security. NPJ Digit Med 2025;8:143. DOI: 10.1038/s41746-025-01476-7.

45. Oh G, Kim S, Park S, Kim BH. Model and task-aware test-time scaling strategies for large language and vision-language models in medicine: evaluation study. J Med Internet Res 2026;28:e90693. DOI: 10.2196/90693.

46. Huang X, Wu J, Liu H, Tang X, Zhou Y. m1: unleash the potential of test-time scaling for medical reasoning with large language models. Proc Mach Learn Res 2026;297:369–383 (https://proceedings.mlr.press/v297/huang26a.html).

